# Remdesivir for the treatment of COVID-19: A living systematic review

**DOI:** 10.1101/2020.09.27.20202754

**Authors:** F Verdugo-Paiva, P Acuña, I Sola, G Rada, COVID-19 L·OVE Working Group

**Affiliations:** Epistemonikos Foundation, Santiago, Chile; UC Evidence Center, Cochrane Chile Associated Center, Pontificia Universidad Católica de Chile, Santiago, Chile; Unidad de Infectología, Hospital Dr Sótero del Río, Santiago, Chile; Unidad de Infectología, Hospital Clínico Dra Eloísa Díaz, La Florida, Santiago, Chile; Biomedical Research Institute Sant Pau, Barcelona, Spain; Iberoamerican Cochrane Centre, Barcelona, Spain; CIBER Epidemiología y Salud Pública (CIBERESP)

**Keywords:** COVID-19, Coronavirus disease, Severe Acute Respiratory Syndrome Coronavirus 2, Coronavirus Infections, Systematic Review, Remdesivir, Antivirals

## Abstract

**Objective:** This living systematic review aims to provide a timely, rigorous and continuously updated summary of the evidence available on the role of remdesivir in the treatment of patients with COVID-19.

**Methods:** We adapted an already published common protocol for multiple parallel systematic reviews to the specificities of this question.

Eligible studies were randomised trials evaluating the effect of remdesivir versus placebo or no treatment.

We conducted searches in the L·OVE (Living OVerview of Evidence) platform for COVID-19, a system that maps PICO questions to a repository maintained through regular searches in electronic databases, preprint servers, trial registries and other resources relevant to COVID-19. All the searches covered the period until 25 August 2020. No date or language restrictions were applied. Two reviewers independently evaluated potentially eligible studies according to predefined selection criteria, and extracted data on study characteristics, methods, outcomes, and risk of bias, using a predesigned, standardised form.

We performed meta-analyses using random-effect models and assessed overall certainty in evidence using the GRADE approach.

A living, web-based version of this review will be openly available during the COVID-19 pandemic. We will resubmit it every time the conclusions change or whenever there are substantial updates.

**Results:** Our search strategy yielded 574 references. Finally, we included 3 randomised trials evaluating remdesivir in addition to standard care versus standard care alone. The evidence is very uncertain about the effect of remdesivir on mortality (RR 0.7, 95% CI 0.46 to 1.05; very low certainty evidence) and the need for invasive mechanical ventilation (RR 0.69, 95% CI 0.39 to 1.24; very low certainty evidence). On the other hand, remdesivir likely results in a large reduction in the incidence of adverse effects in patients with COVID-19 (RR 1.29, 95% CI 0.58 to 2.84; moderate certainty evidence).

**Conclusions:** The evidence is insufficient for the outcomes critical for making decisions about the role of remdesivir in the treatment of patients with COVID-19, so it is not possible to balance the potential benefits, if any, with the adverse effects and costs.

**PROSPERO Registration number:** CRD42020183384

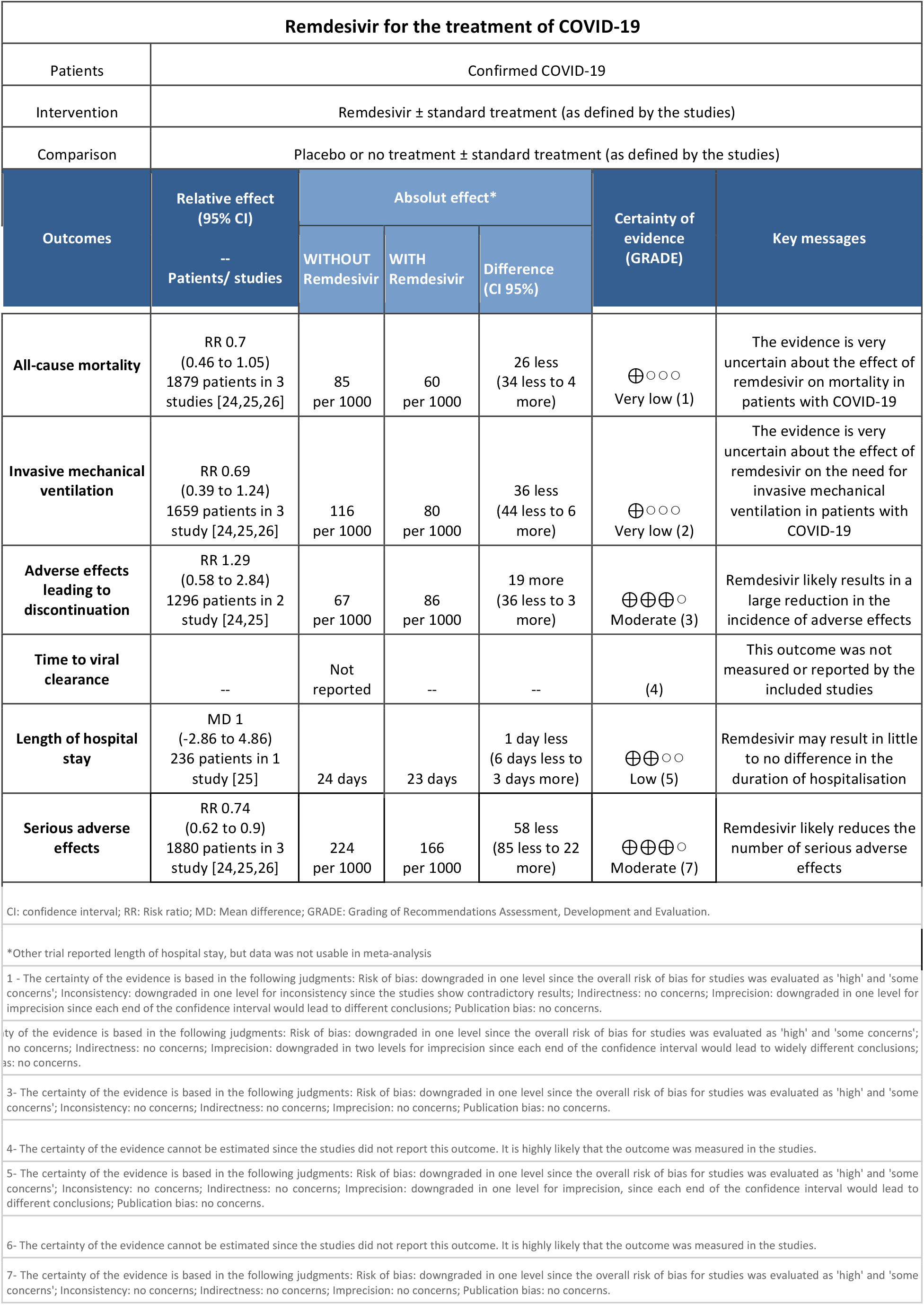

## Introduction

COVID-19 is an infection caused by the SARS-CoV-2 coronavirus [1]. It was first identified in Wuhan, China, on December 31, 2019 [2]; seven months later, more than fifteen million cases of contagion had been identified across 188 countries [3]. On March 11, 2020, WHO characterised the COVID-19 outbreak as a pandemic [1].

While the majority of cases result in mild symptoms, some might progress to pneumonia, acute respiratory distress syndrome and death [4],[5],[6]. The case fatality rate reported across countries, settings and age groups is highly variable, but it ranges from about 0.5% to 10% [7]. In hospitalised patients, it has been reported to be higher than 10% in some centres [8].

One of the strategies underway to identify effective interventions for COVID-19 is repurposing drugs that have been used for the treatment of other diseases.

Remdesivir is among these investigational medications. It is a directly acting antiviral agent, initially developed for the treatment of Ebola virus during the 2014 outbreak in Western Africa [9].

Remdesivir displays antiviral activity against many RNA viruses including SARS-CoV-2, in both in vitro [10] and animal studies [11].

Following the publication of the ACTT-1, a trial conducted by the National Institute of Allergy and Infectious Diseases (NIAID), the US Food and Drug Administration issued an emergency use authorisation of remdesivir for the treatment of COVID-19 [12].

However, the results of ACTT-1 were questioned immediately, particularly for the decision to stop it early for benefit [13]. On the other hand, the decision of the government of the United States of buying virtually all stocks of the drug, generated an urgent need of independent, transparent information about the effects of remdesivir for COVID-19.

Using innovative and agile processes, taking advantage of technological tools, and resorting to the collective effort of several research groups, this living systematic review aims to provide a timely, rigorous and continuously updated summary of the evidence available on the effects of remdesivir in patients with COVID-19.

## Methods

This manuscript complies with the ‘Preferred Reporting Items for Systematic reviews and Meta-Analyses ‘ (PRISMA) guidelines for reporting systematic reviews and meta-analyses [14] (see Appendix 1 - PRISMA Checklist).

A protocol stating the shared objectives and methodology of multiple evidence syntheses (systematic reviews and overviews of systematic reviews) to be conducted in parallel for different questions relevant to COVID-19 was published elsewhere [15]. The review was registered in PROSPERO with the number CRD42020183384 and a full protocol was made available [16].

### Search strategies

#### Electronic searches

We used the search strategies already developed in the L·OVE (Living OVerview of Evidence) platform (https://www.iloveevidence.com), a system that maps the evidence to different research questions. The full methods to maintain L·OVE are described in the website, but the process to devise the search strategies can be briefly described as:

- Identification of terms relevant to the population and intervention components of the search strategy, applying Word2vec technology [17] to the corpus of documents available in Epistemonikos Database.
- Discussion of terms with content and methods experts to identify relevant, irrelevant and missing terms.
- Creation of a sensitive boolean strategy encompassing all the relevant terms
- Iterative analysis of articles missed by the boolean strategy, and refinement of the strategy accordingly.

All the information in the L·OVE platform comes from a repository developed and maintained by Epistemonikos Foundation through the screening of different sources relevant to COVID-19. At the time of releasing this article, this repository included more than 66989 articles relevant to Coronavirus disease, coming from the following databases, trial registries, preprint servers and websites relevant to COVID-19: Epistemonikos database, Pubmed, EMBASE, ICTRP Search Portal, Clinicaltrials.gov, ISRCTN registry, Chinese Clinical Trial Registry, IRCT - Iranian Registry of Clinical Trials, EU Clinical Trials Register: Clinical trials for covid-19, NIPH Clinical Trials Search (Japan) - Japan Primary Registries Network (JPRN) (JapicCTI, JMACCT CTR, jRCT, UMIN CTR), UMIN-CTR - UMIN Clinical Trials Registry, JRCT - Japan Registry of Clinical Trials, JAPIC Clinical Trials Information, Clinical Research Information Service (CRiS), Republic of Korea, ANZCTR - Australian New Zealand Clinical Trials Registry, ReBec - Brazilian Clinical Trials Registry, CTRI - Clinical Trials Registry - India, DRKS - German Clinical Trials Register, LBCTR - Lebanese Clinical Trials Registry, TCTR - Thai Clinical Trials Registry, NTR - The Netherlands National Trial Register,PACTR - Pan African Clinical Trial Registry, REPEC - Peruvian Clinical Trial Registry,SLCTR - Sri Lanka Clinical Trials Registry, medRxiv Preprints, bioRxiv Preprints, SSRN Preprints, WHO COVID-19 database.

The last version of the methods, the total number of sources screened, and a living flow diagram and report of the project is updated regularly on the website [18].

The repository is continuously updated [18] and the information is transmitted in real time to the L·OVE platform, however, it was last checked for this review the day before release on 25 August 2020. The searches covered the period from the inception date of each database, and no study design, publication status or language restriction was applied.

The following strategy was used to retrieve from the repository the articles potentially eligible for this review. coronavir* OR coronovirus* OR betacoronavir* OR “beta-coronavirus” OR “beta-coronaviruses” OR “corona virus” OR “virus corona” OR “corono virus” OR “virus corono” OR hcov* OR covid* OR “2019-ncov” OR cv19* OR “cv-19” OR “cv 19” OR “n-cov” OR ncov* OR (wuhan* and (virus OR viruses OR viral)) OR sars* OR sari OR “severe acute respiratory syndrome” OR mers* OR “middle east respiratory syndrome” OR “middle-east respiratory syndrome” OR “2019-ncov-related” OR “cv-19-related” OR “n-cov-related” AND (remdesivir* OR “GS-5734” OR “GS 5734” OR GS5734*)

### Other sources

In order to identify articles that might have been missed in the electronic searches, we proceeded as follows:

- Screened the reference lists of other systematic reviews.
- Scanned the reference lists of selected guidelines, narrative reviews and other documents.

### Eligibility criteria

We included randomised controlled trials evaluating patients infected with SARS-CoV-2 of any severity.

The intervention of interest was remdesivir at any dosage, duration, timing or route of administration. The comparison of interest was placebo (remdesivir plus standard of care versus placebo plus standard of care) or no treatment (remdesivir plus standard of care versus standard of care).

Our primary outcome of interest was all-cause mortality at longest follow-up. Secondary outcomes were invasive mechanical ventilation and adverse effects leading to discontinuation.

We also extracted information on the following outcomes: time to viral clearance, length of hospital stay and serious adverse effects.

We did not consider the outcomes as an inclusion criteria during the selection process. Any article meeting all the criteria except for the outcome criterion was preliminarily included and assessed in full text.

### Selection of studies

The results of the searches in the individual sources were de-duplicated by an algorithm that compares unique identifiers (database ID, DOI, trial registry ID), and citation details (i.e. author names, journal, year of publication, volume, number, pages, article title, and article abstract). Then, the information matching the search strategy was sent in real-time to the L·OVE platform where at least two authors independently screened the titles and abstracts yielded against the inclusion criteria. We obtained the full reports for all titles that appeared to meet the inclusion criteria or required further analysis and then decided about their inclusion.

We recorded the reasons for excluding trials in any stage of the search and outlined the study selection process in a PRISMA flow diagram which we adapted for the purpose of this project.

### Extraction and management of data

Using standardised forms, two reviewers independently extracted the following data from each included trial: study design, setting, participant characteristics (including disease severity and age) and study eligibility criteria; details about the administered intervention and comparison, including dose, duration and timing (i.e. time after diagnosis); the outcomes assessed and the time they were measured; the source of funding of the study and the conflicts of interest disclosed by the investigators; the risk of bias assessment for each individual study.

We resolved disagreements by discussion, with one arbiter adjudicating unresolved disagreements.

### Risk of bias assessment

The risk of bias for each randomised trial was assessed by using the ‘risk of bias ‘ tool (RoB 2.0: a revised tool to assess risk of bias in randomised trials) [19], considering the following domains of bias for each outcome result of all reported outcomes and time points: bias due to (1) the randomisation process, (2) deviations from intended interventions (effects of assignment to interventions at baseline), (3) missing outcome data, (4) measurement of the outcome, and (5) selection of reported results.

Discrepancies between review authors were resolved by discussion to reach consensus. If necessary, a third review author was consulted to achieve a decision.

### Measures of treatment effect

For dichotomous outcomes, we expressed the estimate of treatment effect of an intervention as risk ratios (RR) along with 95% confidence intervals (CI).

For continuous outcomes, we used the mean difference and standard deviation to summarise the data along with 95% CI. For continuous outcomes reported using different scales, the treatment effect was expressed as a standardised mean difference with 95% CI.

### Strategy for data synthesis

The results of the search and the selection of the studies is presented, by means of the corresponding flow chart, according to recommendations of the PRISMA statement [14]. For any outcomes where it was not possible to calculate an effect estimate, a narrative synthesis is presented, describing the studies in terms of the direction and the size of effects, and any available measure of precision For any outcomes where data was available from more than one trial, we conducted a formal quantitative synthesis (meta-analysis) for studies clinically homogeneous using RevMan 5 [20], using the inverse variance method with the random-effects model. We assessed inconsistency by visual inspection of the forest plots and using the I^2^ index.

### Subgroup and sensitivity analysis

As few trials were found, we did not perform sensitivity or subgroup analysis.

Assessment of certainty of evidence

The certainty of the evidence for all outcomes was judged using the Grading of Recommendations Assessment, Development and Evaluation working group methodology (GRADE Working Group) [21], across the domains of risk of bias, consistency, directness, precision and reporting bias. For the main comparisons and outcomes, we prepared a Summary of Findings (SoF) tables [22],[23].

### Living evidence synthesis

An artificial intelligence algorithm deployed in the Coronavirus/COVID-19 topic of the L·OVE platform provides instant notification of articles with a high likelihood of being eligible. The authors review them, decide upon inclusion, and update the living web version of the review accordingly.

This review is part of a larger project set up to produce multiple parallel systematic reviews relevant to COVID-19 [15].

## Results

### Results of the search

We conducted searches using L·OVE (Living OVerview of Evidence) platform for COVID-19, a system that maps PICO questions to a repository, maintained through regular searches in 27 databases, preprint servers, trial registries and websites relevant to COVID-19. All the searches covered the period until 25 August 2020. No date or language restrictions were applied.

The search in the L·OVE platform yielded 574 records after removal of duplicates. We considered 489 as potentially eligible and obtained and evaluated their full texts. We finally included 3 randomised trials (11 references) [24],[25], [26].

The reasons for excluding studies at the time of full-text review were the following: not a primary study in humans (396 records); wrong study design (51 records) and wrong comparison (3 records). We also identified 16 ongoing randomised trials.

The complete study selection process is summarised in the PRISMA flow chart (Figure 1) and the full list of included, excluded and ongoing trials is presented in Appendix 2.

**Figure 1.**
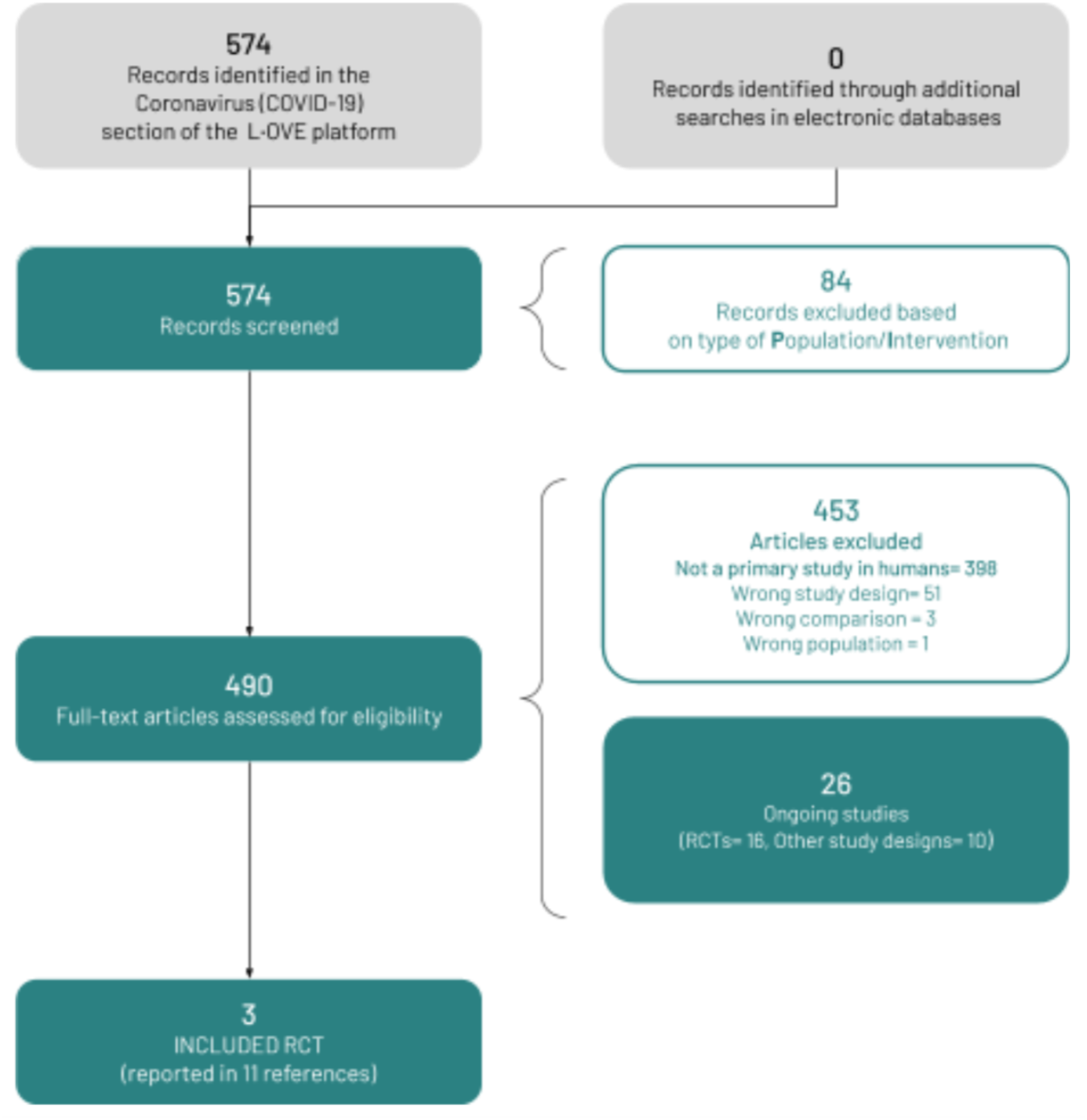
PRISMA Flowchart (prepared by the authors from the study data).

**Figure 2.**
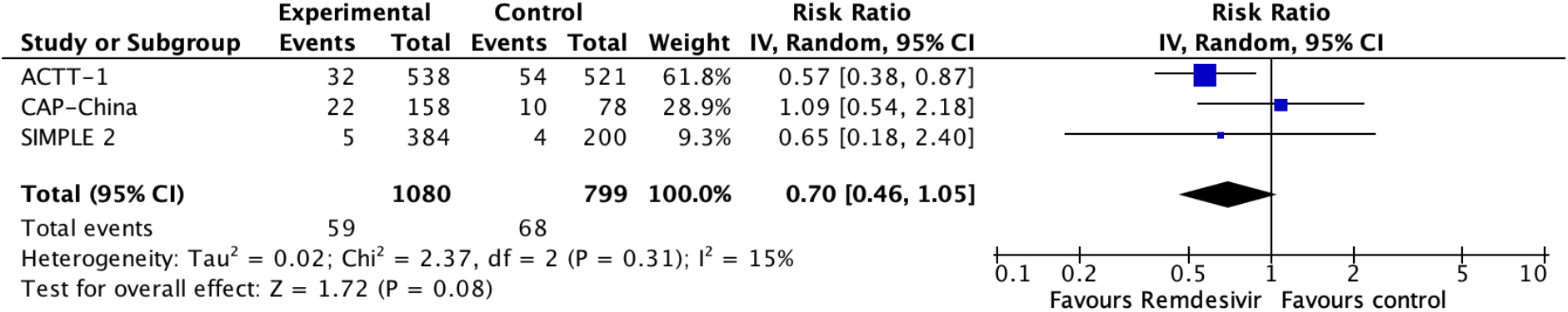
Relative risk for all-cause mortality for remdesivir versus standard care (prepared by the authors from the study data).

**Figure 3.**
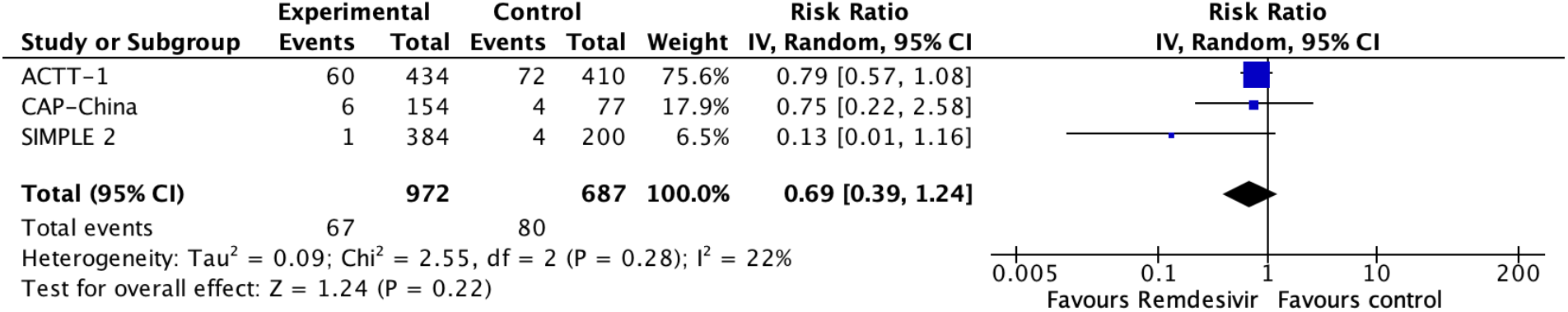
Relative risk for invasive mechanical ventilation for remdesivir versus standard care (prepared by the authors from the study data).

**Figure 4.**
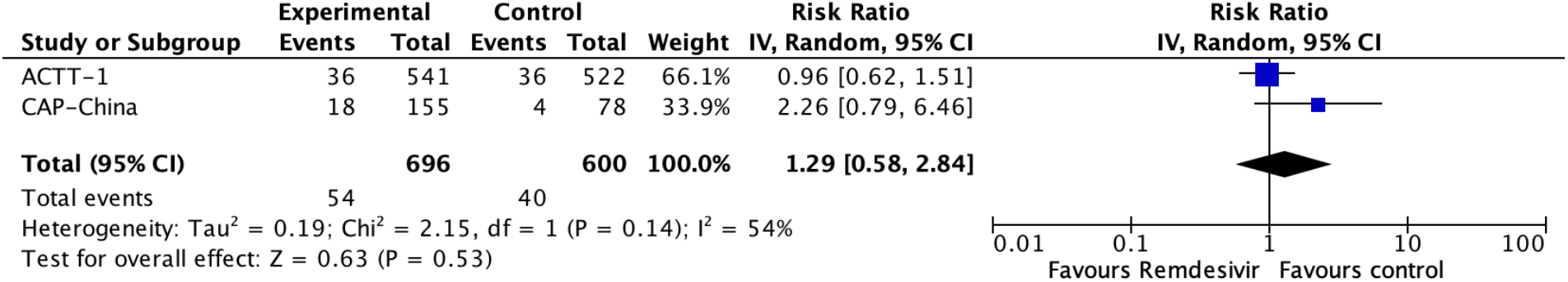
Relative risk for adverse effects leading to discontinuation for remdesivir versus standard care (prepared by the authors from the study data).

**Figure 5.**
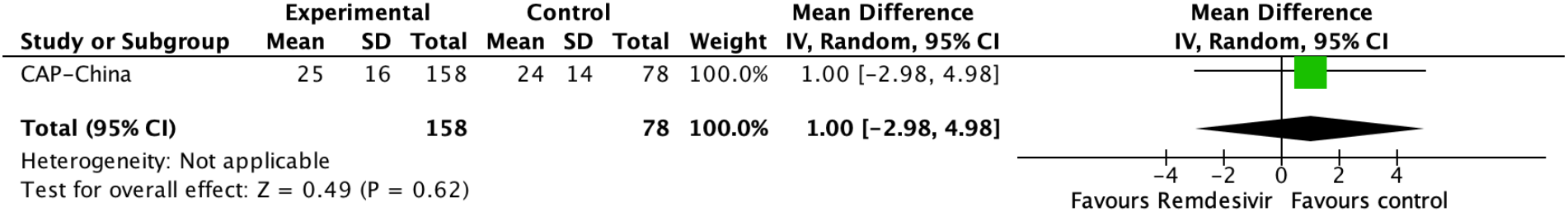
Relative risk for length of hospital stay for remdesivir versus standard care (prepared by the authors from the study data).

**Figure 6.**
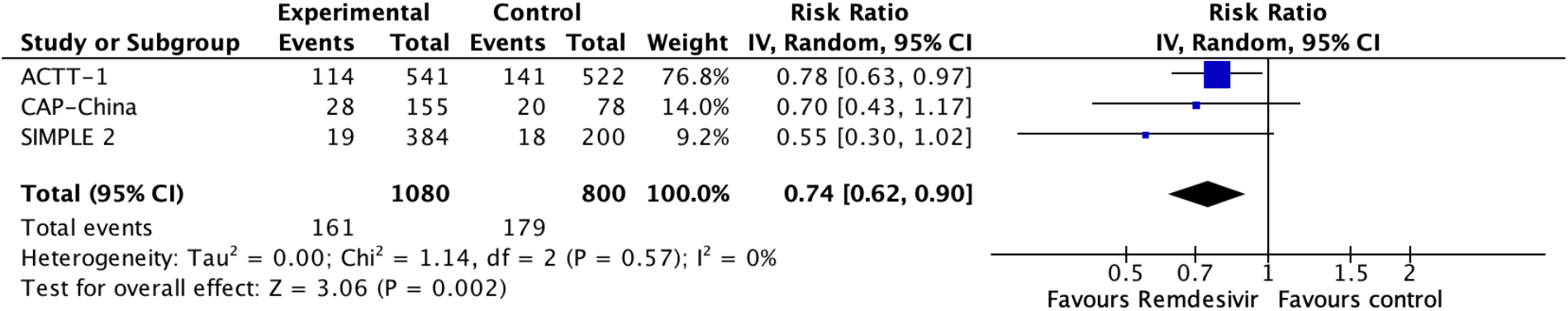
Relative risk for serious adverse effects for remdesivir versus standard care (prepared by the authors from the study data).

### Description of the included studies

The three trials identified were the Adaptive COVID-19 Treatment Trial (ACTT-1 [24]), the CAP-China remdesivir 2 [25] and SIMPLE 2 [26]. All trials evaluated inpatient adults. ACTT-1 required for inclusion that one of the following criteria were also fulfilled: SpO2 </= 94% on room air, requiring supplemental oxygen, requiring mechanical ventilation or radiographic infiltrates by any imaging test. CAP-China remdesivir 2 required that patients had an oxygen saturation of 94% or lower on room air or a ratio of arterial oxygen partial pressure to fractional inspired oxygen of 300 mm Hg or less. Additionally, patients in CAP-China remdesivir 2 had to present within 12 days of symptom onset. SIMPLE 2 [26] required that patients had any radiographic evidence of pulmonary infiltrates and oxygen saturation >94% on room air Table 1 and 2 summarises inclusion criteria of the trials and characteristics of the intervention. More details are presented in Appendix 2. Table 1 presents the complete inclusion criteria of the trials.

**Table 1.** Inclusion criteria of the studies

|  | Age | Setting | Confirmation method | Clinical or severity parameters | Radiological findings as criteria |
| --- | --- | --- | --- | --- | --- |
| ACTT-1 [24] | Adults | Hospital | RT-PCR | One of several criteria (SpO<sub>2</sub> ≤ 94% on room air, OR Requiring supplemental oxygen, OR Requiring mechanical ventilation. ) | One of several criteria (radiographic infiltrates by any imaging test) |
| CAP-China remdesivir 2 [25] | Adults | Hospital | RT-PCR | Mandatory (Oxygen saturation of 94% or lower on room air or a ratio of arterial oxygen partial pressure to fractional inspired oxygen of 300 mm Hg or less, and were within 12 days of symptom onset) | Mandatory (pneumonia confirmed by chest imaging) |
| SIMPLE 2 [26] | Adults and children | Hospital | RT-PCR | Mandatory (SpO<sub>2</sub> > 94% on room air at screening) | Mandatory (any radiographic evidence of pulmonary infiltrates ) |

**Table 2.**
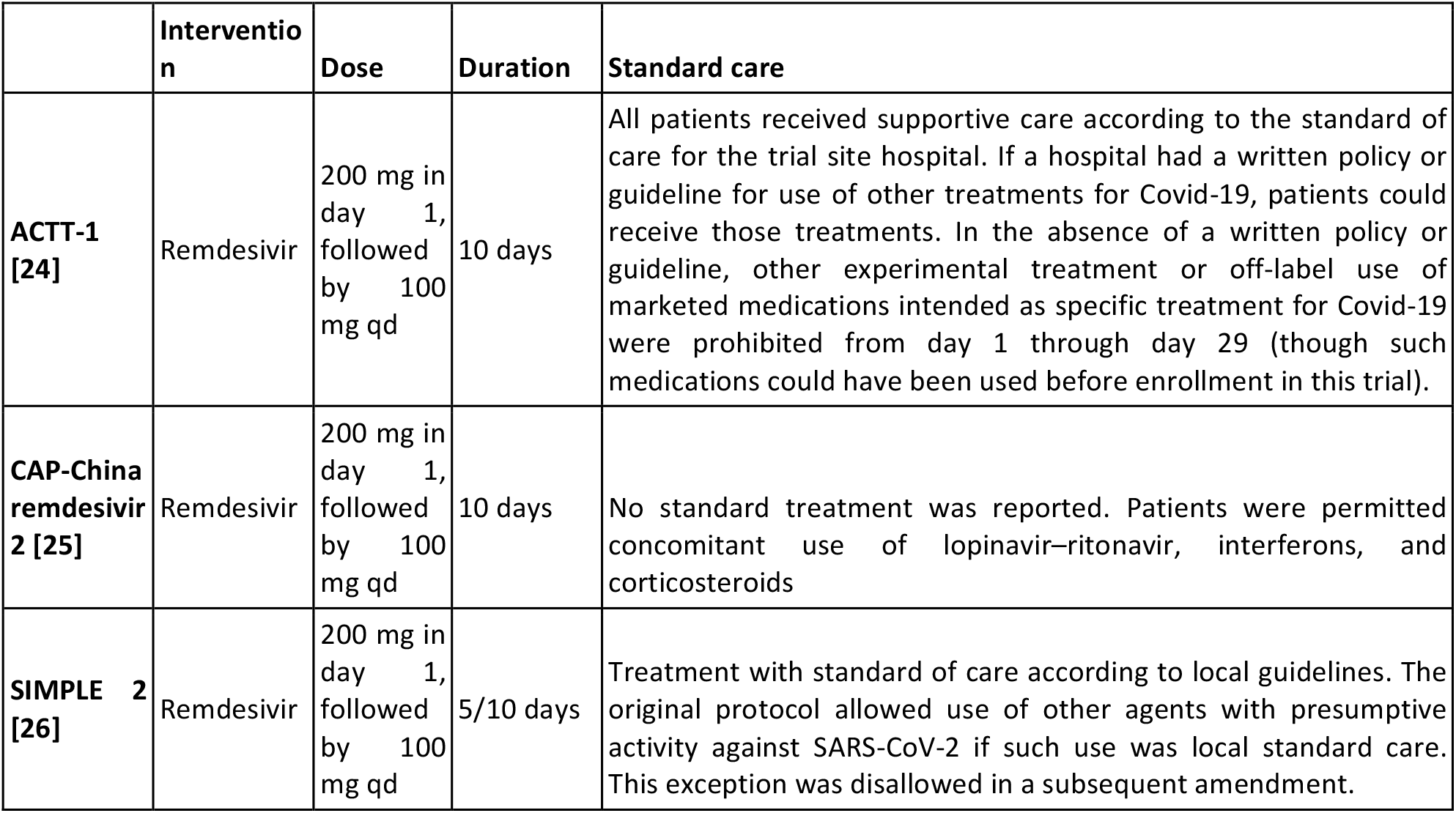
Characteristics of the intervention

All trials administered the same doses of remdesivir plus standard care [24], [25], [26]. One trial included two intervention arms of remdesivir (5-day and 10-day course of remdesivir) [26]. None of the trials provide further details regarding the standard care treatment delivered. Two trials reported that the standard of care was determined by the trial site hospital [24] The other one, only reported that concomitant use of lopinavir/ritonavir, interferons, and corticoids were permitted [25].

In total, trials included 1896 hospitalized patients [24], [25], [26]. One trial was conducted in China [25] and the other two were multicenters trials conducted in several countries [24] [26]. All trials included patients with radiologically confirmed pneumonia [25], [25], [26]. Baseline characteristics of participants regarding age, gender, and chronic disease were similar between studies, but the number of patients requiring supplemental oxygen or mechanical ventilation varied substantially between trials [24], [25].

### Risk of bias in the included studies

We judge that the overall risk of bias was “high” for all outcomes regarding the ACTT-1 trial [24]. The study was judged to raise “some concerns ‘ ‘ in deviations from the intended intervention domain and “high” in bias due to missing outcome data. CAP-China remdesivir 2 trial overall risk of bias was “some concern” for all outcomes, because of problems in the randomization process [25]. SIMPLE 2 overall risk of bias was some concern for all outcomes due to deviations from intended interventions [26]. Table 4 summarises the risk of bias assessments and details of each assessment are presented in Appendix 2.

**Table 3.** Baseline characteristics of the participants

|  | ACTT-1 [24] | CAP-China remdesivir 2 [25] | SIMPLE 2 [26] |
| --- | --- | --- | --- |
| **Number randomised** | 1063 | 237 | 596 |
| **Geographic location and Setting** | United States,Europe and Asia; inpatient setting | China;inpatient setting | United States,Europe and Asia;inpatient setting |
| **Mean age (years)** | 58.9 | 65 | 57 |
| **Females (%)** | 35.7 | 41 | 38.8 |
| **Time from onset to treatment (days)** | 9 | 10 | 9 |
| **Pneumonia (%)** | 100 | 100 | 100 |
| **Supplemental oxygen or NIRS (%)** | 39.6 | 83 | 15.1 |
| **Receiving mechanical ventilation (%)** | 25.6 | 0.3 | Not reported |
| **Underlying chronic diseases (%)** | 49.6% hypertension, 37% obesity, 29.7% diabetes | Hypertension: 72 (46%) vs 30 (38%); Diabetes:40 (25%) vs 16 (21%); Coronary heart disease: 15 (9%) vs 2 (3%) | Cardiovascular disease: 56%, hypertension: 42%, Diabetes 40%, Asthma:14% |

**Table 4.**
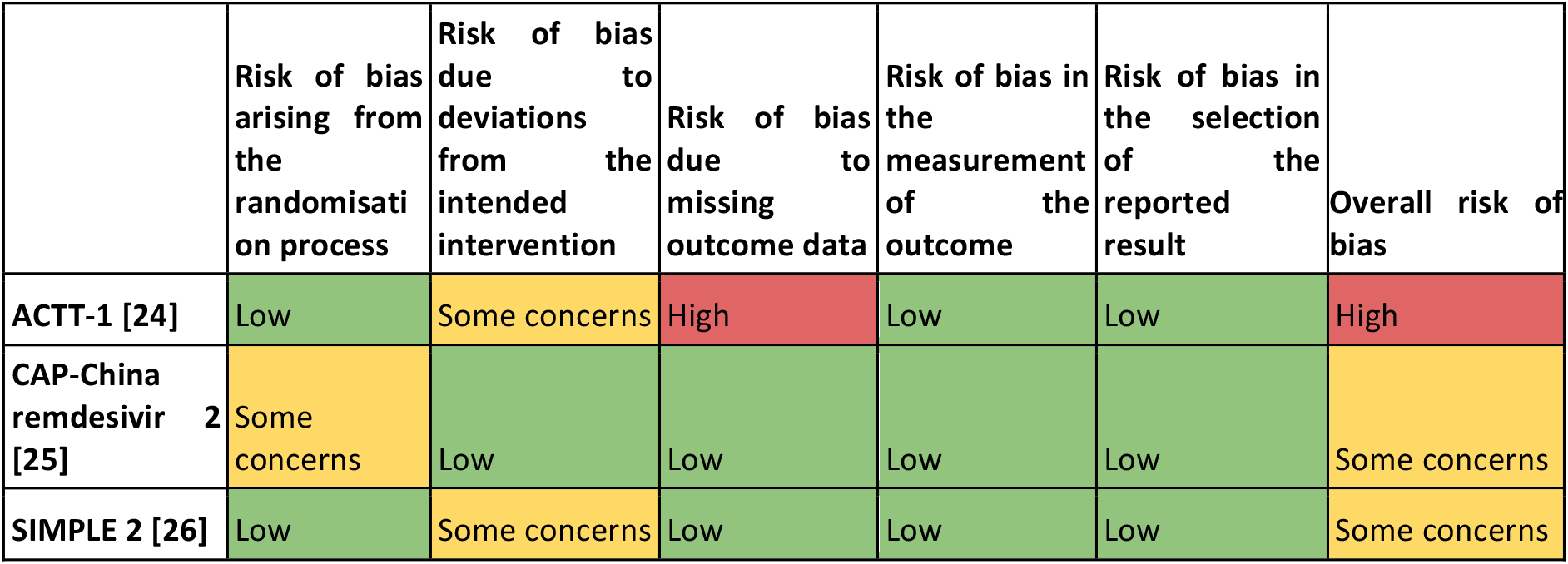
Risk of bias in the included studies assessed by ROB-2 tool

Table 4 summarises the risk of bias assessments and details of each assessment are presented in Appendix 2.

## Efficacy of remdesivir in the treatment of patients with COVID-19

The main results are summarised in the Summary of Findings table, presented at the beginning of the manuscript.

### Primary outcome

#### All-cause mortality

All studies reported this outcome [24],[25],[26] and the evidence is very uncertain about the effect of remdesivir on mortality (RR 0.7, 95% CI 0.46 to 1.05; very low certainty evidence).

### Secondary outcomes

#### Invasive mechanical ventilation

All studies reported this outcome [24],[25],[26] and the evidence is very uncertain about the effect of remdesivir on the need for invasive mechanical ventilation (RR 0.69, 95% CI 0.39 to 1.24; very low certainty evidence).

#### Adverse effects leading to discontinuation

Two trials reported this outcome [24],[25] and remdesivir likely results in a large reduction in the incidence of adverse effects (RR 1.29, 95% CI 0.58 to 2.84; moderate certainty evidence).

#### Time to viral clearance

This outcome was not measured or reported by the included studies

#### Length of hospital stay

Two studies reported this outcome [25],[26], but only one was usable for meta analysis [25]. SIMPLE 2 trial reported that there were no significant differences between the remdesivir and standard care groups in duration of hospitalisation [25]. Quantitative synthesis showed remdesivir may result in little to no difference in the duration of hospitalisation (MD 1, 95% CI −2.86 to 4.86; Low certainty evidence).

### Other outcomes

#### Serious adverse effects

All studies reported this outcome [24],[25],[26] and remdesivir likely reduces the number of serious adverse effects (RR 0.74, 95% CI 0.62 to 0.9; moderate certainty evidence).

## Discussion

We conducted a systematic review and identified 3 randomised trials that reported data on the effect of remdesivir in patients with COVID-19 [24],[25],[26]. Even though remdesivir appears to be safe, the evidence is very uncertain about the impact on the outcomes critical for decision-making in moderate and severe patients, the more relevant clinical scenario for this drug, such as mortality and need of mechanical ventilation.

It is unfortunate not knowing yet if one of the pharmaceutical interventions that has sparked more interest is effective or not. One of the limitations comes from the lack of precision of the result for the main outcomes. The early termination of the ACTT-1 trial can be seen as a missed opportunity in this regard [25].

In addition, all the trials concluded enrollment before the release of the RECOVERY trial which showed a mortality reduction with dexamethasone [27]. It is not clear if this factor would modify the effect, if any, of remdesivir.

By now, clinicians and other decision makers are in a difficult position. The pressure to act is high, particularly after the the US Food and Drug Administration issued an emergency use authorisation of remdesivir for the treatment of COVID-19 [12]. We anticipate that the range of recommendations from different organisations should range between a suggestion against its use and a weak recommendation for its use in severe cases, especially in settings without resource constraints.

There are at least 46 ongoing trials that we expect will provide data in the near future. Making sense of this information is not going to be an easy task. Systematic reviews are considered the gold standard to make sense of multiple trials addressing a similar scientific question, but the traditional model for conducting reviews has several limitations, including a high demand for time and resources [28] and a rapid obsolescence [29]. Amidst the COVID-19 crisis, researchers should make their best effort to answer the urgent needs of health decision makers yet without giving up scientific accuracy. Information is being produced at a vertiginous speed [30], so alternative models are needed.

One potential solution to these shortfalls are rapid reviews, a form of knowledge synthesis that streamlines or omits specific methods of a traditional systematic review in order to move faster. Unfortunately, in many cases, this rapidity comes at the cost of quality [31]. Furthermore, they do not solve the issue of obsolescence. Living systematic reviews do address that issue [32]. They are continually updated by incorporating relevant new evidence as it becomes available, at a substantial effort. So, an approach combining these two models might prove more successful in providing the scientific community and other interested parties with evidence that is actionable, rapidly and efficiently produced, up to date, and of the highest quality [33].

This review is part of a larger project set up to put such an approach into practice. The project aims to produce multiple parallel living systematic reviews relevant to COVID-19 following the higher standards of quality in evidence synthesis production [15]. We believe that our methods are well suited to handle the abundance of evidence that is to come, including evidence on the role of lopinavir/ritonavir for COVID-19.

During the COVID-19 pandemic, we will maintain the search and selection of evidence for this review continuously updated, and we will every time the conclusions change or whenever there are substantial updates. Our systematic review aims to provide high-quality, up-to-date synthesis of the evidence that is useful for clinicians and other decision-makers.

## Data Availability

All data related to the project will be available. Epistemonikos Foundation will grant access to data.

## Notes

## Acknowledgements

The members of the COVID-19 L·OVE Working Group and Epistemonikos Foundation have made possible to build the systems and compile the information needed by this project. Epistemonikos is a collaborative effort, based on the ongoing volunteer work of over a thousand contributors since 2012.

## Roles and contributions

All the review authors drafted and revised the protocol, conducted article screening and data collection, and drafted and revised the review.

The COVID-19 L·OVE Working Group was created by Epistemonikos and a number of expert teams in order to provide decision makers with the best evidence related to COVID-19. Up-to-date information about the group and its member organisations is available here: epistemonikos.cl/working-group

## Competing interests

All authors declare no financial relationships with any organisation that might have a real or perceived interest in this work. There are no other relationships or activities that might have influenced the submitted work.

## Funding

This project was not commissioned by any organisation and did not receive external funding. Epistemonikos Foundation is providing training, support and tools at no cost for all the members of the COVID-19 L·OVE Working Group.

## PROSPERO registration number

CRD42020183384

## Ethics

As researchers will not access information that could lead to the identification of an individual participant, obtaining ethical approval was waived.

## Notes

### Competing Interest Statement

The authors have declared no competing interest.

### Clinical Protocols

https://osf.io/exv6p/

